# Role of T and B lymphocyte cannabinoid type 1 and 2 receptors in major depression and suicidal behaviors: effects of in vitro cannabidiol administration

**DOI:** 10.1101/2023.04.19.23288847

**Authors:** Michael Maes, Muanpetch Rachayon, Ketsupar Jirakran, Atapol Sughondhabirom, Abbas F. Almulla, Pimpayao Sodsai

## Abstract

Early flow cytometry studies revealed T cell activation in major depressive disorder (MDD) (Maes et al., 1990-1993). MDD is characterised by activation of the immune-inflammatory response system (IRS) and the compensatory immunoregulatory system (CIRS), including deficits in T regulatory (Treg) cells. This study examines the number of cannabinoid type 1 (CB1) and type 2 (CB2) receptor bearing T/B lymphocytes in MDD, and the effects of in vitro cannabidiol (CBD) administration on CB1/CB2. Using flow cytometry, we determined the percentage of CD20+CB2+, CD3+CB2+, CD4+CB2+, CD8+CB2 and FoxP3+CB1+ cells in 19 healthy controls and 29 MDD patients in 5 conditions: baseline, stimulation with anti-CD3/CD28 with or without 0.1 µg/mL, 1.0 µg/mL or 10.0 µg/mL CBD. We found that CB2+ was significantly higher in CD20+ than CD3+ and CD4+, and CD8+ cells. Stimulation with anti-CD3/CD8 beads increases the number of CB2-bearing CD3+, CD4+, and CD8+ cells, as well as CB1-bearing FoxP3+ cells. There was an inverse association between the number of reduced CD4+CB2+ and IRS profiles, including M1 macrophage, T helper-(Th)-1 and Th-17 phenotypes. MDD is characterized by lowered basal FoxP3+CB1+% and higher CD20+CB2+%. 33.2% of the variance in the depression phenome (including severity of depression, anxiety, and current suicidal behaviors) is explained by CD20+CB2+% (positively) and CD3+CB2+% (inversely). All 5 immune cell populations were significantly increased by 10 µg/mL CBD administration. In conclusion, reductions in FoxP3+CB1+% and CD3+/CD4+CB2+% contribute to deficits in immune homeostasis in MDD, while increased CD20+CB2+% may contribute to the pathophysiology of MDD by activating T-independent humoral immunity.

**Summations:**

- Lowered CD4+CB2+ T cells are associated with increased immune-inflammatory responses (IRS) in major depressive disorder (MDD)
- Lowered CD3+CB2+% and increased CD20+CB2+% predict severity of depression and suicidal behaviors
- Lowered CD3/CD4+CB2+ may impact the immune homeostatic processes leading to enhanced IRS in MDD
- Increased CD20+CB2+% may activate T-independent humoral immunity and enhance IRS responses.

**Considerations:**

- Depletion of CB1 bearing T regulatory and CB2 bearing T and T helper cells and increased CB2+ bearing B cells are new drug targets in MDD.
- The findings deserve replication in other countries and cultures.
- Future research should examine CB2 bearing macrophages, dendritic cells, and natural killer cells in MDD

## Introduction

There is now evidence that major depressive disorder (MDD) is a neuro-immune disorder characterised by multiple immune system dysfunctions that culminate in an increase in neuronal and astroglial toxicity (Maes, 1995; 2022; Maes and Carvalho, 2018; Al-Hakeim et al., 2023). Initial studies demonstrated that MDD is associated with elevated levels of pro-inflammatory cytokines of the M1 macrophage lineage, such as interleukin (IL)-1β, IL-1 receptor antagonist (sIL-1RA), IL-6, IL-8 (or CXCL8), and tumour necrosis factor (TNF)-α, and the T helper (Th)-1 lineage, such as IL-2, IL-2 receptor (sIL-2R), and interferon (IFN)-γ (Maes et al., 1990; 1991; 1994; 1995b; Mikova et al., 2001). Additional research conducted in the laboratories of Maes et al. demonstrated elevated levels of complement factors and positive acute phase proteins, such as haptoglobin, and decreased levels of negative acute phase proteins, such as albumin, in MDD, indicating that MDD is accompanied by a chronic mild inflammatory („acute phase") response (Maes, 1993). The cytokine, macrophage-T lymphocyte, and immune-response system (IRS) theories of MDD summarised these findings (Maes, 1995; Maes et al., 1995a). Recent systematic reviews and meta-analyses corroborate that MDD is associated with elevated serum levels of M1 macrophage and Th1 cytokines and immune activation (Kohler et al., 2017; Zhang et al., 2023; Foley et al., 2023; Gasparini et al., 2022; Mac Giollahui et al., 2021).

Recent research indicates that MDD is characterised not only by activation of M1, Th-1, and Th-17 activation but also the compensatory immunoregulatory system (CIRS), which prevents hyperinflammation and endeavours for immune homeostasis (Maes et al., 2012; Maes and Carvalho, 2018). Th-2 (producing IL-4) and T regulatory (Treg, producing IL-10) phenotypes are major CIRS components that are activated in MDD (Maes et al., 2022). In MDD, T cell growth factors such as IL-4, IL-9, IL-12, and granulocyte-macrophage colony-stimulating factor (GM-CSF) contribute to the simultaneous activation of both IRS and CIRS components (Maes et al., 2022). Importantly, certain IRS and CIRS cytokines (IL-1β, IL-2, IL-4, IL-6, TNF-α, IFN-γ) and chemokines (CCL1, CCL2, CCL11, CXCL5, CXCL8, CXCL10) have neurotoxic effects and could, therefore, contribute to the neuronal and astroglial projection toxicity in MDD (Maes et al., 2022; Al-Hakeim et al., 2023).

Early machine learning and flow cytometry studies to quantify the percentage and number of peripheral blood mononuclear cells (PBMC) revealed IRS activation, particularly T cell activation, in patients with MDD with an increased expression of CD4+, CD45+, CD25+, Ig+, and HLA-DR+ on peripheral blood mononuclear cell (PBMC) (Maes et al., 1990; 1992a; 1992b; 1993). With a specificity of 92%, machine learning revealed that up to 64% of MDD patients exhibited elevated expression of CD7+CD25+ and CD2+HLADR+ cells (Maes et al., 1993). T regulatory (Treg) cells are also involved in the pathophysiology of MDD. For example, MDD patients have lower Treg levels than healthy controls and in animal models lowered Treg function is associated with depression-like behaviors (Grosse-Hoogenboezem et al., 2016; Kim et al., 2012). A decrease in T effector cells and an increase in Treg expression characterise the symptomatic remission phase of a major depressive episode of bipolar disorder (Maes et al., 2021). Antidepressant-treated depressed patients had a greater number of CD4+CD25+ and CD4+CD25+FOXP3+Treg than untreated depressed patients (Mohd-Ashari et al., 2019). Treg cells, such as CD4+CD25+FoxP3+, CD4+CD25+FoxP3+CD152+, and CD4+CD25+FoxP3+GARP+, are essential for immune tolerance and homeostasis, the suppression of T cell activation, and immune proliferation (Maes et al., 2021).

B cells and monocytes express higher levels of CB2 receptors than CD4+ (T helper) and CD8+ (T cytotoxic) T lymphocytes (Graham et al., 2010). Dendritic cells and macrophages may release endogenous cannabinoids in response to inflammatory stimuli, whereas activating CB2 receptors has profound anti-inflammatory effects by inhibiting T cell activation, M1 macrophage functions, immune cell proliferation and differentiation, and inducing apoptosis in both T and B cells (Turcotte et al., 2016; Ehrhart et al., 2005; Klein et al., 2004; Komorowska-Muller et al., 2020; Morcuende et al., 2022; Csoka et al., 2009; Lombard et al., 2007; Rieder et al., 2009; Maresz et al., 2007). When CB2 is activated, M1 macrophages acquire a M2 phenotype and produce more IL-10, an important CIRS cytokine (Correa et al., 2005; Klein et al., 1998). Transgenic mice overexpressing CB2 (CB2xP) show reduced depressive-like behaviors and are resistant to acute anxiogenic stimuli (Garcia-Gutierrez et al., 2023).

CB2 is crucial for the homing, development, and maintenance of marginal zone (MZ) B cells and enhances T-cell-independent humoral responses of natural IgM (Basu et al., 2011; 2013; Nolte et al., 2004). These IgM are crucial as the first line innate immune defense against pathogens, including commensal microorganisms (Zouali and Richard, 2011; Morris et al., 2019). This is significant because MDD patients have elevated IgM antibody levels against a variety of endogenous antigens and LPS of gut commensal bacteria as well (Maes et al., 2011; 2013; Simeonova et al., 2021). In this respect, there are some data that CB2 may contribute to bacterial translocation via increased production of IL-10 (Csoka et al., 2009). CB1 is also expressed on immune cells and, like CB2 and also in conjunction with CB2, displays immunosuppressive effects, such as suppression of Th-1 functions (Kaplan et al., 2013).However, there are no data on the number of CB2/CB1-bearing CD3+ (pan T), CD4+, CD8+, CD20+ (pan B), and FoxP3+ cells in patients with MDD compared to healthy controls.

Cannabidiol (CBD), a phytocannabinoid extracted from the cannabis plant, is a potential antidepressant (Atalay et al., 2019). There is evidence that CBD may improve clinical depression and clinical anxiety (Martin et al., 2021), although not all studies show positive results (Hasbi et al., 2023). Some, but not all, animal models of depression show that cannabinoids have antidepressive effects (Hen-Shoval et al., 2018; Hasbi et al., 2023). CBD has a lower affinity for CB2 receptors than anandamide (AEA), 2-arachidonoylglycerol (2-AG), and delta-9-tetrahydrocannabinol (THC) (Pertwee et al., 2007; Raich et al., 2021), but possesses anti-inflammatory and antioxidative properties, such as by inhibiting the production of pro-inflammatory cytokines and reactive oxygen species (Atalay et al., 2019). CBD is a partial agonist at the CB2 receptor that modifies the efficacy of cannabinoid ligands, suggesting that CBD’s anti-inflammatory and atidepressant effects may be related to their effects on CB2 (An et al., 2020). However, there are no data indicating whether CBD may affect the number or percentage of CB2/CB1 bearing CD3+, CD4+, CD8+, CD20+, and FoxP3+ cells.

In light of this, the current study examined the number of CB2/CB1-containing CD3+, CD4+, CD8+, CD20+, and FoxP3+CB1+ cells in patients with MDD compared to healthy controls, as well as the in vitro effects of CBD on these cell populations.

## Methodology

### Participants

We recruited outpatients with MDD admitted to the psychiatry department of the Chulalongkorn Hospital in Bangkok, Thailand, for this study. Healthy controls were recruited from the same geographic region (Bangkok, Thailand) as the patients via posters and word of mouth. The inclusion criteria were patients between the ages of 18 and 65 who spoke Thai, had been diagnosed with MDD by a senior psychiatrist using DSM-5 criteria, and had a Hamilton Depression Rating Scale (HAM-D) score of at least 17. Excluded were healthy participants with a diagnosis of any axis 1 DSM-5 disorder or a positive family history of MDD or bipolar disorder (BD). Patients with BD, schizophrenia, substance use disorders, schizoaffective disorders, psycho-organic disorders, obsessive-compulsive disorder, and post-traumatic stress disorder were excluded from the study. Exclusion criteria for both patients and controls were: neuroinflammatory and neurodegenerative disorders such as multiple sclerosis, Alzheimer’s disease, epilepsy, or Parkinson’s disease; (auto)immune illnesses such as inflammatory bowel disease, chronic obstructive pulmonary disease, cancer, diabetes type 1, asthma, psoriasis, or rheumatoid arthritis; and allergic or inflammatory responses in the past three months. Participants who took anti-inflammatory medications (NSAIDs or steroids) within one month of the study, therapeutic doses of antioxidant or omega-3 polyunsaturated fatty acid supplements within three months prior to the study, or those who had ever received immunomodulatory drugs were also excluded. Neither lactating women nor pregnant women were allowed to participate.

Some patients were taking psychotropic medications, such as sertraline (18 patients), other antidepressants (8 patients, such as fluoxetine, venlafaxine, escitalopram, bupropion, and mirtazapine), benzodiazepines (22 patients), atypical antipsychotics (14 patients), and mood stabilizers (4 patients). In the statistical analysis, the possible effects of these pharmacological variables were taken into consideration.

Before participating in the investigation, all subjects and controls provided written consent. The research was conducted in accordance with both international and Thai privacy and ethics regulations. The Institutional Review Board of the Faculty of Medicine at Chulalongkorn University in Bangkok, Thailand approved the study (#528/63) in accordance with international guidelines for the protection of human research subjects, including the Declaration of Helsinki, The Belmont Report, CIOMS Guideline, and the International Conference on Harmonization in Good Clinical Practice (ICH-GCP).

### Clinical measurements

A well-trained research assistant conducted semi-structured interviews to collect demographic data, including age, gender, medical and psychiatric history. The 17-item HAMD was administered by an experienced psychiatrist to determine the severity of depression (Hamilton, 1960). The Mini-International Neuropsychiatric Interview (M.I.N.I.) was used to diagnose axis-1 psychiatric illness (Kittirattanapaiboon and Khamwongpin, 2005). The Thai state version of the State-Trait Anxiety Inventory (STAI) was used to assess the level of self-rated state anxiety (Spielberger, 1983). Suicidal ideation and behavior were evaluated using the lifeline version of the Columbia-Suicide Severity Rating Scale (C-SSRS) (Posner et al., 2011). Recent suicidal behaviors were computed as the first principal component (PC) (labelled "Current_SB") extracted from 9 C-SSRS items: “wish to be dead, non-specific active suicidal thoughts, active suicidal ideation with any methods, active suicidal ideation with some intent to act, active suicidal ideation with a specific plan/intent, frequency and duration of suicidal ideation, actual attempts, and the total number of actual attempts". This first PC extracted from the 9 SB items possessed satisfactory psychometric properties (Maes et al., 2022). The “phenome of depression” was calculated by extracting the first PC from the total HAMD, STAI, and Current_SB scores (Maes et al., 2022). This PC exhibited excellent psychometric properties and loadings > 0.90 on all indicators. Body mass index (BMI) was calculated by dividing body weight (in kilograms) by length squared (in meters squared). Using DSM-5 criteria, a tobacco use disorder (TUD) was diagnosed.

### Assays

Fasting blood samples of 20 mL were collected between 8-10 a.m. in BD Vacutainer® EDTA (10 mL) and BD Vacutainer® SST™ (5 mL) tubes (BD Biosciences, Franklin Lakes, NJ, USA). To obtain serum, blood was allowed to clot at room temperature for 30 minutes. The tubes were spun at 1100 ×g for 10 minutes at 4°C, and the resulting serum or plasma was stored at - 80°C. PBMCs were separated through density gradient centrifugation (30 min at 900 ×g) using Ficoll® Paque Plus (GE Healthcare Life Sciences, Pittsburgh, PA, USA). Cell count and viability were checked using hemocytometer and trypan blue, 0.4% solution, pH 7.2-7.3 (Sigma-Aldrich Corporation, St. Louis, MO, USA). The number of total and blue staining cells were counted, ensuring that more than 95% of cells were viable for all conditions.

To evaluate PBMC stimulation, the 96-well plates were coated with 5 µg/mL of the anti-human CD3 antibody (OKT3, from eBioscience), overnight. PBMCs, 3 x 10^5^ cells each, were added to each well, along with 5 μg/mL of the anti-human CD28 antibody (CD28.2, eBioscience). The cells were grown in a culture medium of RPMI 1640 with L-glutamine that was supplemented with 10% fetal bovine serum and 1% penicillin-streptomycin solution (Gibco Life Technologies, Carlsbad, CA, USA). The cells were allowed to grow for 3 days at 37° C in an environment with 5% CO_2_. A negative control was conducted by growing PBMCs without anti-CD3 and anti-CD28 antibodies for the same period. After 3 days, the immunophenotypes of CD20, CD3, CD4, CD8, and Treg cells as well as CB2/CB1 receptors were determined through flow cytometry. Thus, the specimens can be divided into two conditions: unstimulated and stimulated with anti-CD3 and anti-CD28, after being incubated for 72 hours. The effects of three CBD concentrations on the stimulated immune cells were examined using CBD 0.1 µg/ml (CBDlow), 1 µg/mL (CBDmedium), and 10 µg/mL (CBDhigh) (Rachayon et al., 2022). The natural CBD (99.89% isolate) stock preparation was manufactured by Love Hemp Ltd, United Kingdom (batch 8406), and the Medical Cannabis Research Institute, College of Pharmacy, RSU, Bangkok, Thailand, guaranteed its quality. Concentrations between 0.1 – 1.0 µg/mL are therapeutical concentrations of CBD, whereas levels as high as 5-20 µg/mL may be obtained when combining oral CBD coupled with dietary lipids (Rachayon et al., 2022).

To study lymphocyte phenotypes, 3 x 10^5^ PBMCs were labeled with a combination of monoclonal antibodies for 30 minutes, including CD3-PEcy7, CD4-APCcy7, CD8-APC, CD20-FITC and CB2-AF700 antibodies (Biolegend). **Electronic Supplementary File (ESF1), Figure 1** shows our FoxP3+CB1+ gating strategy. We first stained PBMCs with surface markers CD3-PEcy7, CD4-APCcy7, CD25-APC, 7-AAD, and CB1-AF700 (human CB1 receptor) (Biolegend). Intracellular Foxp3 staining was performed using the FoxP3/Transcription Factor Staining Buffer Set (eBioscience) for fixation and permeabilization and following staining with antibody to Foxp3-FITC (Biolegend). Flow cytometry was performed directly after staining, and at least 50,000 lymphocytes were identified based on size and granularity. ESF, Figure 2 shows our gating strategy to assay CD4/CD8+/CD20+CB2+ cells. The flow cytometry was performed using an LSRII flow cytometer (BD Biosciences), and data were analyzed using FlowJo X software (Tree Star Inc., Ashland, OR, USA). As such, we determined the percentage of 5 different phenotypes, namely CD20+CB2+, CD3+CB2+, CD4+CB2+, CD8+CB2+, and FoxP3+CB1+ cells and this in 5 conditions: unstimulated or baseline, anti-CD3 and anti-CD28 stimulated, stimulated + 0.1 µg/mL CBD, stimulated + 1.0 µg/mL CBD, and stimulated + 10.0 µg/mL CBD.

**Figures 1.**
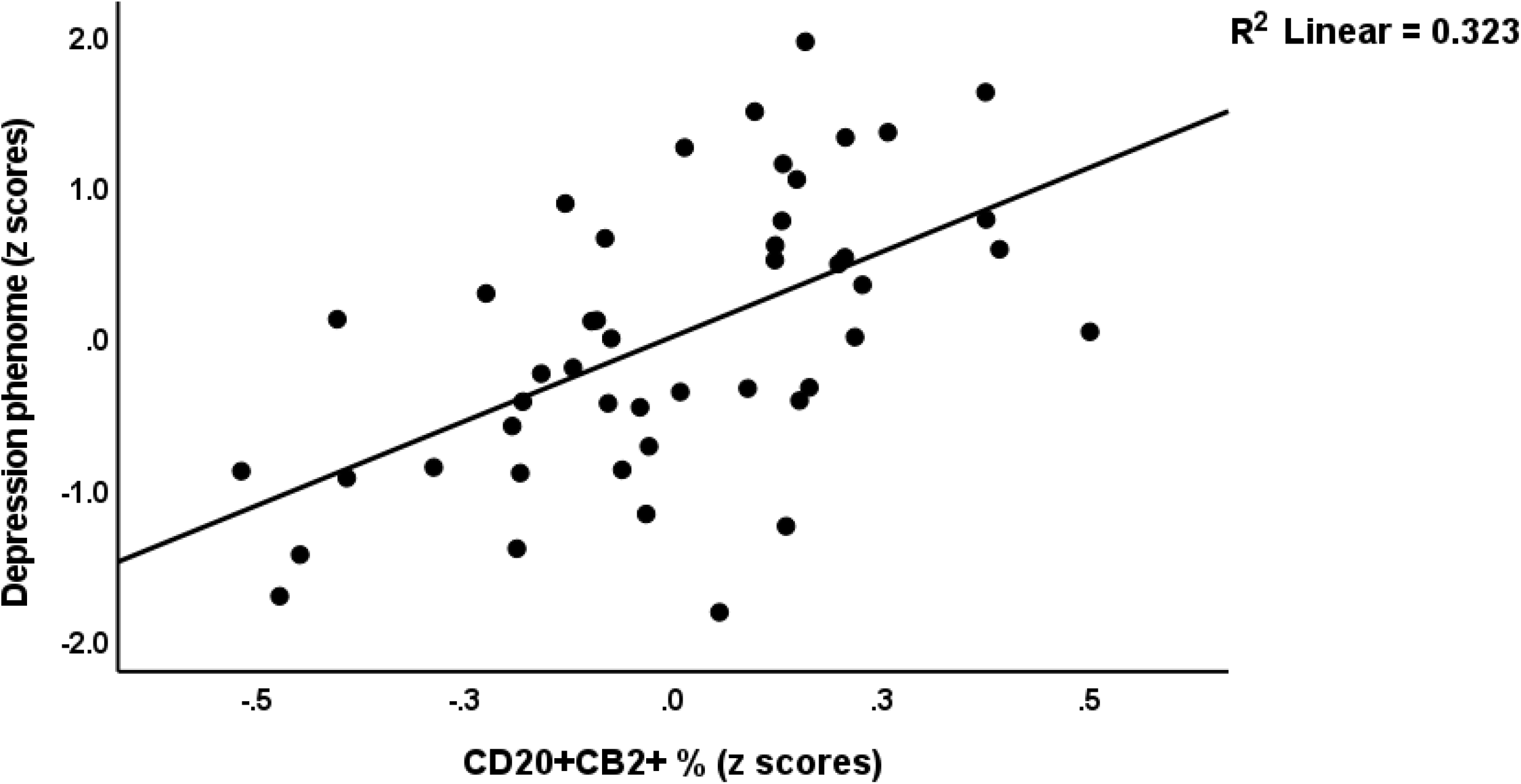
Partial regression of the depression phenome on stimulated CD20+CB2+ cells.

Using the same blood samples we also quantified cytokines/chemokines in stimulated diluted whole blood culture supernatant, as explained previously (Maes et al., 2022). We utilized RPMI-1640 medium supplemented with L-glutamine and phenol red and containing 1% penicillin (Gibco Life Technologies, USA), with or without 5 µg/mL PHA (Merck, Germany) + 25 µg/mL lipopolysaccharide (LPS; Merck, Germany). On 24-well sterile plates, 1.8 mL of each of these two mediums was mixed to 0.2 mL of 1/10 diluted whole blood. The specimens from each individual were incubated for 72 hours at 37°C, 5% CO2 in a humidified atmosphere. After incubation, the plates were centrifuged at 1500 rpm for 8 minutes. Supernatants were carefully extracted under sterile conditions, divided into Eppendorf tubes, and immediately frozen at -70°C until thawed for cytokine/chemokines/growth factor assays. The cytokines/chemokines/growth factors were measured using the Bio-Plex Pro human cytokine 27-plex assay kit (BioRad, Carlsbad, California, United States of America) using the LUMINEX 200 equipment (BioRad, Carlsbad, California, United States of America). For all investigations, the intra-assay CV values are less than 11%. **ESF, Table 1** shows the immune profiles used in the present study.

### Statistics

We compared nominal variables using analysis of contingency tables (χ2-test) and continuous variables using analysis of covariance (ANCOVA) among research groups. For the purpose of examining the connections between scale variables, we employed Pearson’s product moment correlation coefficients, shown in correlograms (heatmap correlation matrix). A first pre-specified generalized estimating equation (GEE), repeated measures, was used to assess the responsivity of the cell populations to administration of anti-CD3/CD28 beads (unstimulated versus stimulated condition), the differences between patients and controls, and cell types (CD3, CD4, CD8, CD20) and all interactions, while allowing for the effects of age, sex, BMI, smoking, and drug state of the patients. A second pre-specified GEE, repeated measures, performed on the stimulated cell population only, included fixed categorical effects of diagnosis, treatment (4 conditions: anti-CD3/CD28 with and without the three CBD concentrations), cell type, and their interactions, while allowing for the effects of the above confounding variables. It should be stressed that there were no missing variables in any of the cell population or clinical data. Manual multiple regression analysis was used to assess the effects of explanatory variables (cell populations) on dependent variables (clinical profiles). In order to decide which variables would be included and which would be excluded in the final regression model, we also used an automatic forward stepwise regression strategy with a p-to-enter of 0.05, and a p-to-remove of 0.06. We estimated the standardized coefficients with t-statistics and exact p-values, F statistics (and p values), and total variance used as effect size (partial Eta squared). We investigated the likelihood of multicollinearity and collinearity using the tolerance (cut-off value: 0.25) and variance inflation factor (cut-off value: > 4), as well as the condition index and variance proportions from the collinearity diagnostics table. The modified Breusch-Pagan test and the White test were used to investigate possible heteroskedasticity. In the final model, the residuals, residual plots, and data quality were always examined. Using the outcomes of the linear modeling analyses, we also computed partial regression analyses, including partial regression plots. The primary statistical analysis is the multiple regression analysis that delineates the prediction of the depression phenome on the immune cell populations. To normalize the data distribution of the indicators, we employed transformations such as logarithmic, square-root, rank-based inversed normal (RINT), and Winsorization when necessary. All tests were two tailed at p=0.05. We utilized IBM’s Windows version of SPSS 28 to conduct the aforementioned statistical analyses. A priori power analysis (G*Power 3.1.9.4) for a linear multiple regression shows that given an effect size of 0.25 (equivalent to 20% explained variance), power=0.8, alpha=0.05 and 3 covariates, the minimum sample size should be 48.

## Results

### Sociodemographic and clinical data

**Table 1** shows that there are no differences in age, sex, and years of education among the study groups. BMI was somewhat higher in patients, but covarying for BMI in the different analyses did not show any effect of BMI on the immune data. The HAM-D, STAI, Current_SB, and phenome scores were significantly higher in patients as compared with controls. Table 1 also shows that the M1, IRS and Tcell growth profiles were significantly increased in MDD, while there was a trend toward increased Th-1 and Th-17 scores.

**Table 1.** Clinical and immune features of heathy controls (HC) and major depressed patients (MDD).

| Features | HC<br>N=19 | MDD<br>N=29 | F / $\chi^2$ | df | p |
| --- | --- | --- | --- | --- | --- |
| Age (years) | 33.8 ± 8.2 | 28.9 ± 8.61 | 3.94 | 1/46 | 0.053 |
| Sex (M/F) | 6/13 | 11/18 | 0.20 | 1 | 0.653 |
| Education (years) | 16.0 ± 2.3 | 15.7 ± 1.4 | 0.53 | 1/46 | 0.473 |
| Body mass index (kg/m<sup>2</sup>) | 21.46 ± 2.5 | 25.0 ± 6.0 | 6.19 | 1/46 | 0.017 |
| HAMD | 0.95 ± 1.58 | 23.4 ± 5.83 | MWUT | - | <0.001 |
| STAI | 37.8 ± 10.9 | 56.7 ± 7.3 | MWUT | - | <0.001 |
| Current_SB (z scores) | -0.915 ± 0.00 | 0.563 ± 0.836 | MWUT | - | <0.001 |
| Phenome (z scores) | -1.135 ± 0.255 | 0.759 ± 0.416 | MWUT | - | <0.001 |
| M1 (z scores) | -0.404 ± 0.273 | 0.264 ± 1.203 | 5.65 | 1/46 | 0.022 |
| Th-1 (z scores) | -0.307 ± 0.537 | 0.201 ± 0.177 | 3.10 | 1/46 | 0.085 |
| Th-17 (z scores) | -0.325 ± 0.520 | 0.213 ± 1.117 | 3.50 | 1/46 | 0.068 |
| IRS (z scores) | -0.415 ± 0.318 | 0.272 ± 1.191 | 5.99 | 1/46 | 0.018 |
| T cell growth (z scores) | -0.409 ± 0.294 | 0.268 ± 1.199 | 5.81 | 1/46 | 0.020 |

All results are shown as mean (SD) or as frequencies, F : results of ANOVA, $\chi^2$ : results of analysis of contingency tables, MWUT: Mann-Whitney U test, HAMD: Hamilton Depression Rating Scale, STAI: State-Trait Anxiety Inventory, Current\_SB: current suicidal behaviors, Phenome: a score based on the HAMD, STAI and Current\_SB scores, M1: macrophage M1, Th: T helper, IRS: immune-inflammatory response system.

### Baseline and stimulated CB2 and CB1 frequencies in controls and patients

GEE analysis performed on the frequency of CB2-bearing cells in the unstimulated and stimulated conditions showed significant effects of stimulation (Wald=7.37, df=1, p=0.007), cell type (Wald or W=60.75, df=3, p<0.001), diagnosis X cell type (W=15.42, df=4, p=0.004), and stimulation X cell type (17.41, df=3, p<0.001). All GEE analyses concerning the CB2+/CB1+ data were adjusted for the possible effects of in vivo treatment with psychotropic drugs, however, no significant effects could be found. **Table 2** shows the significant interaction pattern between stimulation X cell type. After stimulation, the CB2+ percentage on CD3+ (p<0.001), CD4+ (P<0.001) and CD8+ (p=0.003), but not CD20+ (p=0.110) cells, had increased. In the baseline and stimulated conditions, the percentage of CB2 bearing cells decreased from CD20+ → CD3+ and CD4+ → CD8+ (p<0.001). GEE analysis performed on the CD25+FOXP3+CB1+ percentage showed a significant stimulating effect of time (W=9.33, df=1, p=0.002), as shown in Table 2. In baseline conditions, the CD25+FOXP3+CB1+ percentage was lower in MDD than in controls (p=0.042), whereas after stimulation no differences could be found (p=0.902).

**Table 2.** Differences in CB2 and CB1-bearing cells between the unstimulated (baseline) and anti-CD3/CD28-stimulated condition

| Cell type | Baseline condition<br>n=48 | Anti-CD3/CD28<br>stimulated condition<br>n=48 | Pairwise comparison (p-value) |
| --- | --- | --- | --- |
| CD20+CB2+% | 5.38 ± 1.01 | 7.74 ± 1.07 | 0.110 |
| CD3+CB2+% | 1.39 ± 0.29 | 3.22 ± 0.47 | <0.001 |
| CD4+CB2+% | 1.24 ± 0.22 | 3.28 ± 0.52 | <0.002 |
| CD8+CB2+% | 0.91 ± 0.20 | 1.45 ± 0.28 | 0.003 |
| FoxP3+CB1+% | 1.70 ± 0.53 | 3.51 ± 0.26 | 0.002 |

Results of generalized estimating equation, repeated measures, indicating a significant interaction between cell type and stimulation by antiCD3/CB28..

**Table 3** shows the significant interaction between diagnosis X 4 cell types (CD20+, CD3+, CD4+ and CD8+). GEE analysis performed on the baseline and stimulated conditions showed that there were significant effects of diagnosis (W=4.58, df=1, p=0.032) and interaction diagnosis X stimulation (W=3.83, df-1, p=0.050). The percentage of CD20+CB2+ was significantly higher in patients with MDD (8.15 ±1.05%) compared with controls (4.98 ±1.06%), whereas there were no significant differences in CD3+CB2+, CD4+CB2+ and CD8+CB2+ cells between MDD and controls. The interaction pattern showed that the percentage of CD20+CB2+ was higher in the stimulated MDD groups than in the three other groups.

**Table 3.** The significant interaction pattern between diagnosis and cell type in patients with major depressive disorder (MDD) and healthy controls (HC).

| Cell type | HC | MDD | Pairwise comparison (p-value) |
| --- | --- | --- | --- |
| CD20+CB2+% | 4.98 ±1.06 | 8.15 ±1.04 | 0.016 |
| CD3+CB2+% | 2.37 ±0.40 | 2.25 ±0.42 | 0.837 |
| CD4+CB2+% | 2.26 ±0.37 | 2.27 ±0.47 | 0.986 |
| CD8+CB2+% | 1.44 ±0.42 | 0.92 ±1.15 | 0.191 |

Results of generalized estimating equation.

### Regression analyses of severity of illness on CB2 percentages

Consequently, we have examined whether the different immune populations and the immune profiles (M1, Th-1, Th-17, IRS, and T cell growth) predict the clinical scores. **Table 4** shows the results of these multiple regression analyses of clinical severity scores on the unstimulated and stimulated percentages of the immune cells with and without the immune profiles. Regression #1 shows that 29.6% of the variance in the HAMD score was explained by the regression on basal FoxP3+CB2+% and education (both inversely) and stimulated CD20+CB2+% (positively). Regression #2 shows that 34.8% of the variance in the total STAI score was explained by FoxP3+CB2+% and basal CD3+CB2+% (both inversely) and stimulated CD20+CB2+% (positively). Regression #3 shows that 21.2% of the variance in Current_SB was explained by the stimulated values of CD20+CB2+% (positively). Regression #4 shows that 33.2% of the variance in the phenome score was explained by the regression on stimulated values of CD20+CB2+% (positively) and CD3+CB2+%. **Figures 1 and 2** show the partial regression of the phenome on CD20+CB2+% and CD3+CB2+%, respectively.

**Table 4.**
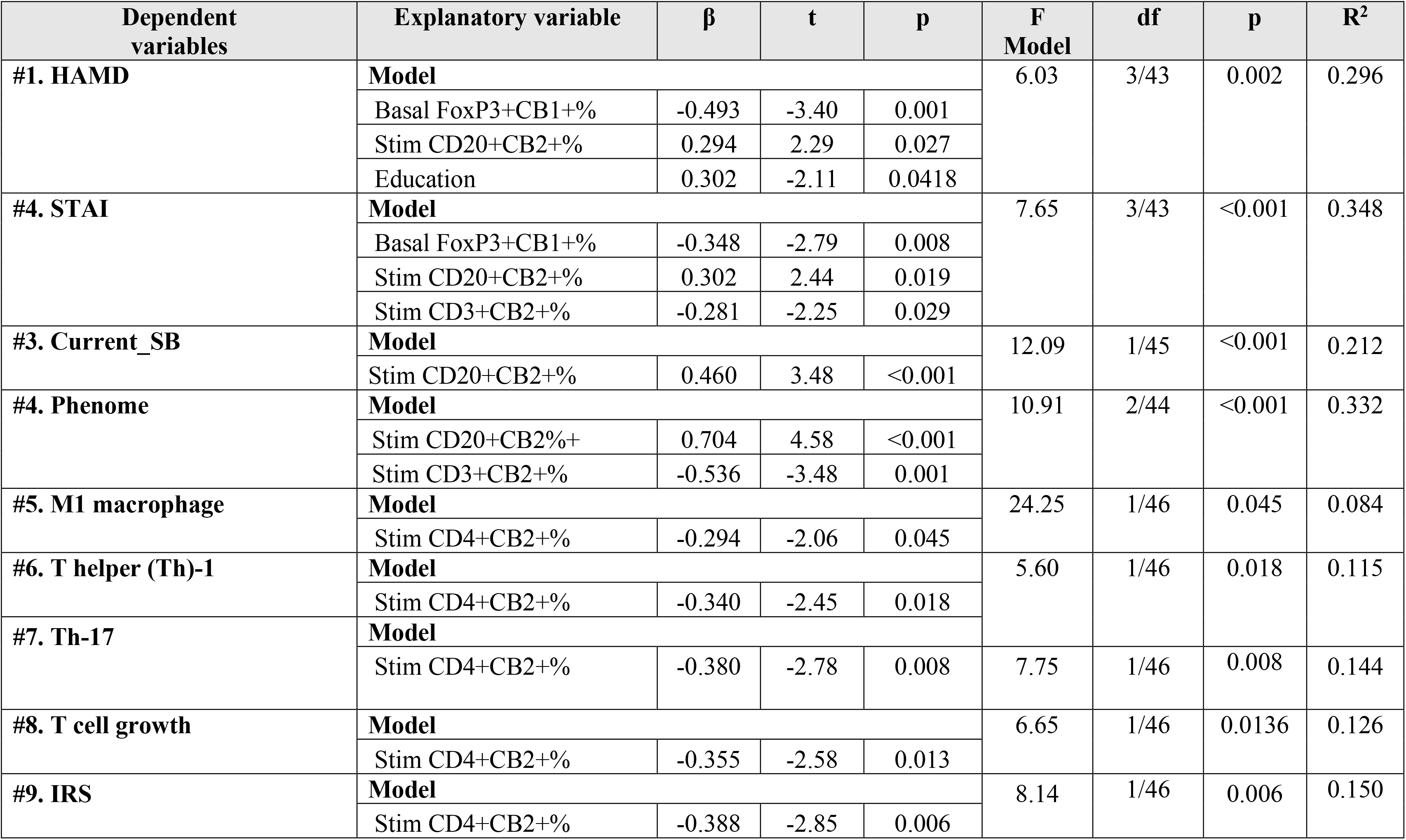
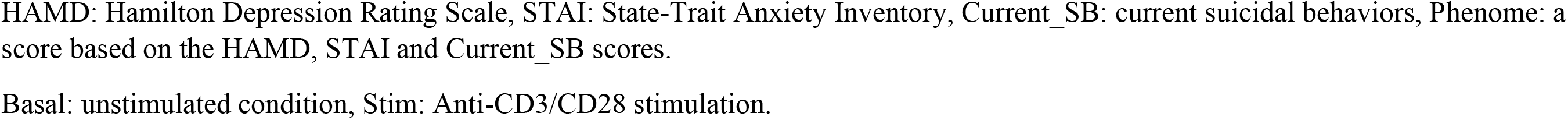
Results of multiple regression analyses with clinical scores or immune profiles as dependent variables and percentage of CB2 and CB1 bearing lymphocytes as explanatory variables.

| Dependent variables | Explanatory variable | $\beta$ | t | p | F Model | df | p | R<sup>2</sup> |
| --- | --- | --- | --- | --- | --- | --- | --- | --- |
| #1. HAMD | **Model** |  |  |  | 6.03 | 3/43 | 0.002 | 0.296 |
|  | Basal FoxP3+CB1+% | -0.493 | -3.40 | 0.001 |  |  |  |  |
|  | Stim CD20+CB2+% | 0.294 | 2.29 | 0.027 |  |  |  |  |
|  | Education | 0.302 | -2.11 | 0.0418 |  |  |  |  |
| #4. STAI | **Model** |  |  |  | 7.65 | 3/43 | <0.001 | 0.348 |
|  | Basal FoxP3+CB1+% | -0.348 | -2.79 | 0.008 |  |  |  |  |
|  | Stim CD20+CB2+% | 0.302 | 2.44 | 0.019 |  |  |  |  |
|  | Stim CD3+CB2+% | -0.281 | -2.25 | 0.029 |  |  |  |  |
| #3. Current_SB | **Model** |  |  |  | 12.09 | 1/45 | <0.001 | 0.212 |
|  | Stim CD20+CB2+% | 0.460 | 3.48 | <0.001 |  |  |  |  |
| #4. Phenome | **Model** |  |  |  | 10.91 | 2/44 | <0.001 | 0.332 |
|  | Stim CD20+CB2+% | 0.704 | 4.58 | <0.001 |  |  |  |  |
|  | Stim CD3+CB2+% | -0.536 | -3.48 | 0.001 |  |  |  |  |
| #5. M1 macrophage | **Model** |  |  |  | 24.25 | 1/46 | 0.045 | 0.084 |
|  | Stim CD4+CB2+% | -0.294 | -2.06 | 0.045 |  |  |  |  |
| #6. T helper (Th)-1 | **Model** |  |  |  | 5.60 | 1/46 | 0.018 | 0.115 |
|  | Stim CD4+CB2+% | -0.340 | -2.45 | 0.018 |  |  |  |  |
| #7. Th-17 | **Model** |  |  |  | 7.75 | 1/46 | 0.008 | 0.144 |
|  | Stim CD4+CB2+% | -0.380 | -2.78 | 0.008 |  |  |  |  |
| #8. T cell growth | **Model** |  |  |  | 6.65 | 1/46 | 0.0136 | 0.126 |
|  | Stim CD4+CB2+% | -0.355 | -2.58 | 0.013 |  |  |  |  |
| #9. IRS | **Model** |  |  |  | 8.14 | 1/46 | 0.006 | 0.150 |
|  | Stim CD4+CB2+% | -0.388 | -2.85 | 0.006 |  |  |  |  |

HAMD: Hamilton Depression Rating Scale, STAI: State-Trait Anxiety Inventory, Current\_SB: current suicidal behaviors, Phenome: a score based on the HAMD, STAI and Current\_SB scores.
Basal: unstimulated condition, Stim: Anti-CD3/CD28 stimulation.

**Figures 2.**
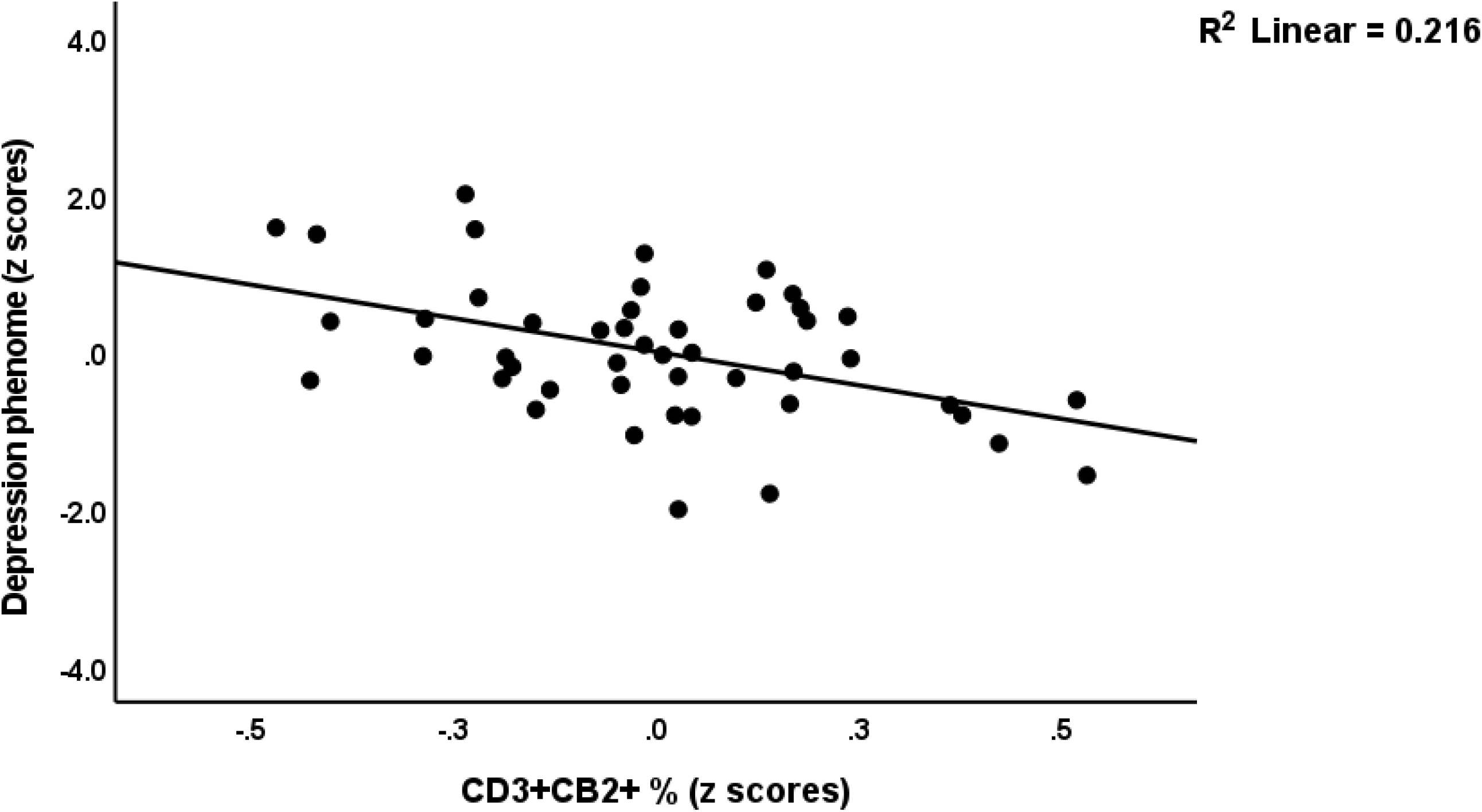
Partial regression of the depression phenome on baseline CD3+CB2+ cells.

We also examined whether the immune profiles could be predicted by the immune cell populations and found that CD4+CB2+% was the single best significant predictor (inversely) of M1 (explained variance: 8.4%), Th-1 (11.5%), Th-17 (14.4%), IRS (11.0%) and T cell growth (11.9%). **Figure 3** shows the regression of T cell growth on CD4+CB2+%.

**Figure 3.**
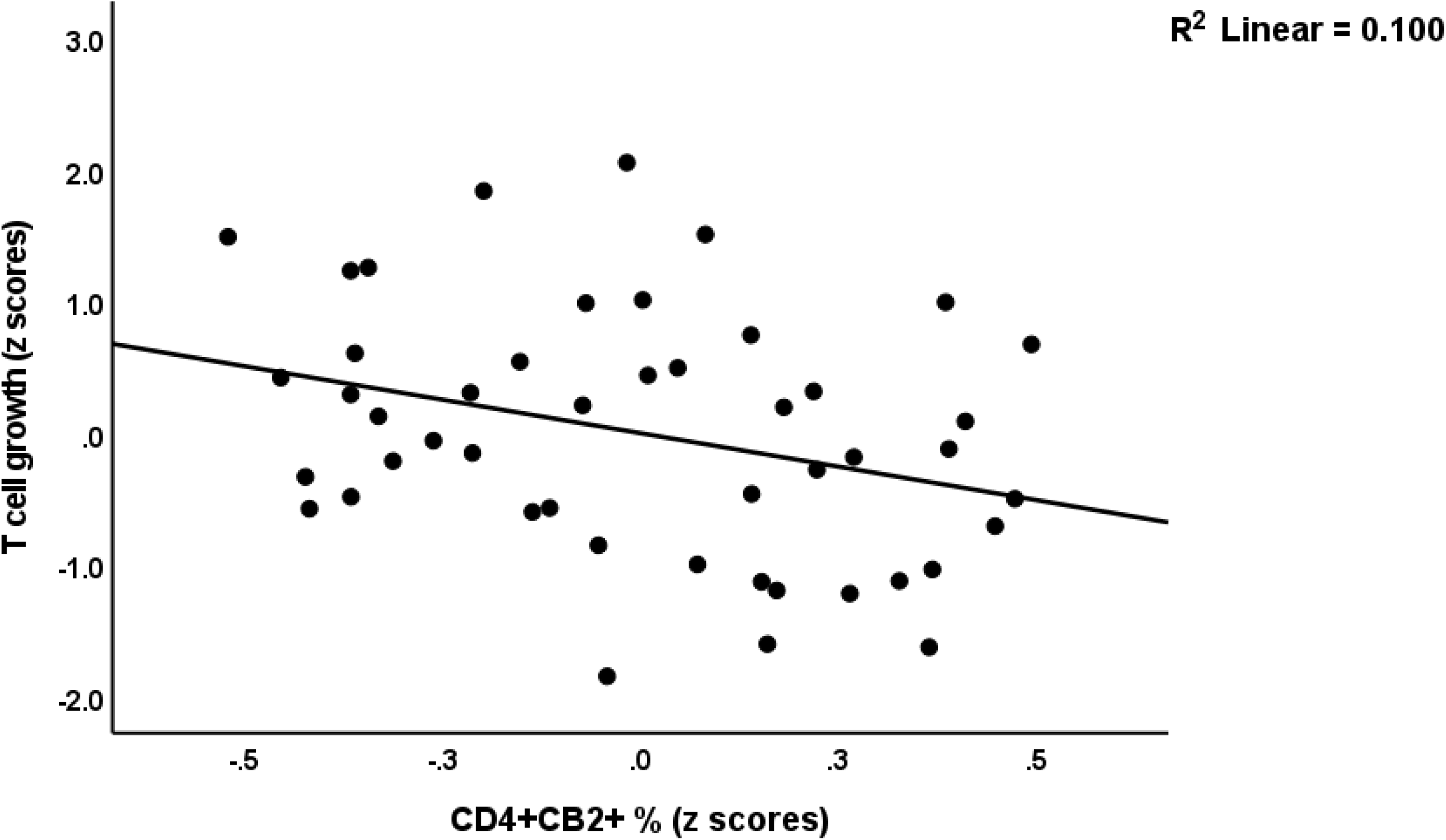
Partial regression of T cell growth on CD4+CB2+ cells.

### Effects of in vitro administration of CBD

GEE analysis performed on all subjects with stimulated conditions (including the 3 CBD conditions) showed significant effects of the stimulation/CBD treatment condition (Wald=18.59, df=3, p<0.001), cell type (W=156.34, df=3, p<0.001), diagnosis (W=4.00, df=1, p<0.045), diagnosis X cell type (W=37.05, df=3, p<0.001), stimulation/CBD treatment X cell type (W=21.48, df=3, p<0.001) on the frequency of CB2-bearing cells. **Table 5** shows the mean percentage of CB2 bearing cells in the stimulated and 3 CBD conditions. The results show that CBDlow and CBDmedium had no significant effect above and beyond anti-CD3/CD28, whereas CBDhigh significantly enhances all 4 cell types highly significantly (p<0.001).

**Table 5.** . Mean percentage of cannabinoid type 2 receptor (CB2) bearing cells in the anti-CD3/CD28 stimulated condition and three cannabidiol (CBD) treatment conditions.

| Estimated marginal means (SE) |  |  | 95% Wald confidence interval |  |
| --- | --- | --- | --- | --- |
| Treatment | Mean | SE | Lower | Upper |
| Anti-CD3/CD28 | 2.586 | .539 | 1.531 | 3.642 |
| Anti-CD3/CD28 + CBD 0.1 µg/dL | 2.858 | .574 | 1.733 | 3.982 |
| Anti-CD3/CD28 + CBD 1 µ/mL | 2.619 | .554 | 1.533 | 3.705 |
| Anti-CD3/CD28 + CBD 10 µg/mL | 8.956 | 1.324 | 6.362 | 11.550 |

Results of generalized estimating equation.

**ESF, Table 2** shows the effects of the three concentrations of CBD on the anti-CD3/CD28 stimulated FoxP3CB1 percentage. GEE analysis showed a significant effect of treatment (W=18.87, df=3, p<0.001) but no significant effects of diagnosis, diagnosis X treatment, and use of in vivo psychotropic drugs. Pairwise comparisons show that the FoxP3+CB1+ percentage was significantly increased by 10 µg/mL CBD as compared with the other 3 stimulated conditions (p<0.001).

## Discussion

### CB2/CB1 bearing T and B lymphocytes

The first major finding of this study is that the CB2% was significantly higher in CD20+ cells than CD3+ and CD4+ cells, and again higher than in CD8+ cells; stimulation with anti-CD3/CD8 beads increases the number of CB2-bearing CD3+, CD4+, and CD8+ cells, as well as CB1-bearing FoxP3+ cells; and lower levels of CD4+CB2+ cells are predictive of increased M1, Th-1, Th-17, IRS, and T cell growth profiles.

CB2 receptors are predominantly expressed in the immune system and the central nervous system (CNS) (including hippocampal neurons and glial cells), whereas CB1 receptors are predominantly expressed in the CNS, although immune cells also express CB1 receptors (Bie et al., 2018; Cabral and Griffin-Thomas, 2009). In fact, previous research has demonstrated that B cells express the greatest quantity of CB2 protein and mRNA, followed by natural killer cells, macrophages, and T cells (Galiegue et al., 1995; Schatz et al., 1997; Lee et al., 2002). Additionally, there is evidence that immune activation is accompanied by increased CB2 expression on tonsillar B cells via CD40 activation (Lee et al., 2002) and that during inflammation, there are more CB2 molecules available for stimulation (Cabral and Griffin-Thomas, 2009). Increased CB2 expression in the brain occurs in response to inflammatory conditions and is predominantly localised in activated microglia, but not in quiescent microglia (Nunez et al., 2004; Cabral and Marciano-Cabral, 2005; Fernandez-Ruiz et al., 2007). Importantly, peripherally activated immune cells that infiltrate the CNS via blood-brain barrier breakdown may contribute to neuroinflammation (Cabral et al., 2008). Stimulation of immune cells, including with antibodies, may increase CB1 mRNA in murine whole spleen cells, whereas stimulation with anti-CD3 antibodies may decrease CB1 mRNA (Noe et al., 2002).

In our study, decreased levels of CD4+CB2+ cells predicted increased immune-inflammatory profiles. This is consistent with previous research demonstrating that increased CB2 expression or B2 ligation maintains immune homeostasis, and has anti-proliferative and anti-inflammatory effects by downregulating T cell functions and T cell effector activities, Th-17 activation, CD4+ cell proliferation, IFN-γ and IL-2 production, macrophage and chemotaxis, and inducing Th-2 polarisation (Ziring D et al., 2006; Cabral and Griffin-Thomas, 2009; Gentili et al., 2019; Eisenstein and Meissler, 2015; Guillot et al., 2014). It should be noted that activated CD20+CB2+ cells may increase IL-10, a negative immunoregulatory cytokine, and are implicated in Th-2 polarisation (Carter et al., 2012; Saroz et al., 2019; Agudelo et al., 2008). On the basis of these findings, CB2 ligation with selective agonists is deemed an effective treatment for conditions marked by immune activation and autoimmunity (Eisenstein and Meissler, 2015). CB2 on autoreactive T cells, for instance, may suppress autoimmune inflammation in the CNS (Maresz et al., 2007). Immune CB1 may also possess anti-inflammatory and immunosuppressive properties, inhibit the production of pro-inflammatory cytokines such as IL-2 and IL-12, and induce apoptosis in immune cells (Kaplan, 2013).

### Lowered immune homeostasis in MDD

The second main finding of this study is that MDD is associated with decreased baseline levels of FoxP3+CB1+ cells and that decreased FoxP3+CB1+% and CD3+CB2+% strongly predicted the phenome of depression, including the severity of depression, anxiety and current suicidal behaviors. Reduced FoxP3+ and CD3+CB2+ are likely to reduce the immune homeostatic processes of CB2/CB1 on T cells and FoxP3+ cells, thereby reducing the immunosuppressive and anti-inflammatory capacities of CB2 and CB1. Consequently, deficiencies in CB receptor markers may account for at least some of the immune findings in depression, including increased T cell activation, CD4+ cell proliferation, increased IL-2 and IFN-γ production, Th-1 polarisation, Th-17 activation, and decreased Treg differentiation (Maes et al., 1990; Maes et al., 2022). Moreover, the cell population markers evaluated in this study are better predictors of the phenome of depression than the immune profiles, which we computed using pro-inflammatory cytokines, chemokines, and T cell growth factors. Interestingly, LPS, which is elevated in MDD due to leaky gut and increased bacterial translocation (Maes et al., 2013), may downregulate CB2 in splenocytes (Lee et al., 2002). Therefore, it is safe to hypothesize that the increased bacterial translocation in MDD may have impacted CB2%. The consequent decreased immune homeostasis as a result of deficits of not only CD3+CB2+% and CD4+CB2+%, but also FoxP3+CB1+% may, therefore, contribute to the peripheral inflammatory and neuroinflammatory responses, which contribute to the pathogenesis of severe MDD.

### Increased CD20+CB2+ cells in MDD

The third main finding of this study is that MDD is associated with a strong increase in CD20+CB2+% following anti-CD3/CD28 activation, and that this is markedly and positively associated with the depression phenome and recent suicidal behaviors. In significant subgroups of persons with MDD, previous research has reported both T cell activation and B cell proliferation (Maes, 1995). Importantly, we found that patients with MDD have a substantially increased percentage and expression of CD154+ (CDL40+) on CD3+ and CD4+ cells, indicating T cell activation via the T cell receptor (TCR) complex, which is a component of CD3 and CD4/8 (Rachayon et al., submitted). T cell activation via anti-CD3/CD28 beads involves three steps: a) TCR engagement with the beads (or antibodies by antigen-presenting cells); b) co-stimulatory signals through CD28, which further activate CD4+ and CD8+ T cells; and c) T-dependent activation of B cells via CD40-CD40L interactions (Thermo-Fisher, 2023; Grewal and Flavell, 1996; Klaus et al., 1994). In fact, the upregulation of CD40L is a strict requirement for T-dependent B cell activation (Klaus et al., 1994). Moreover, CD40 ligation results in the upregulation of CB2 on B cells, with CB2+ B cells playing an essential role in B cell differentiation (Carayon et al., 1998). These findings indicate that the increased proportion of CB2+ B cells in MDD is likely the result of T cell activation and, more specifically, CD40-CD40L interactions.

However, CB2 is also a crucial component of T-independent humoral immunity and B cell-associated immunity (Basu et al., 2011). Importantly, CB2 upregulates MZ B cells, which, as part of the innate immune response, produce natural IgM (see Introduction; Basu et al., 2011; Rayman et al., 2004). Recent reviews demonstrate that the polyreactive natural IgM produced by B1 cells is directed against a number of self-antigens and commensal bacterial antigens, exerts protective, anti-inflammatory, and antioxidative effects, clears cell debris, and serves as a first-line defense against pathogen invasion (Morris et al., 2019; Allman et al., 2019). CB2 deficiency induces deficits in natural IgM and humoral immune responses (Basu et al., 2011). In addition, T cell-independent antigens, such as certain commensal microbes and viruses, may prime MZ B cells to respond quickly to activation stimuli (Basu et al., 2011). As discussed in the Introduction, MDD is associated with elevated levels of IgM antibodies to numerous self-antigens, oxidatively and nitrosatively modified neoepitopes, and the LPS of commensal Gram-negative bacteria. Consequently, T cell activation and the resulting increase in CD20+CB2+% may play a crucial role in the increase in natural IgM levels.

Nevertheless, these B cells can also have detrimental effects, such as pro-inflammatory effects, activating innate response activator cells, acting as antigen-presenting cells that stimulate T cells to produce more pro-inflammatory cytokines, inducing Th-1 and Th-17 responses, and inhibiting Treg cell formation (Morris et al., 2019). In addition, some B1 cells produce high affinity IgM which activates the complement system and induces inflammation (Kerfoot et al., 2008; Askenase et al., 2015). Abnormal B1 cell trafficking may contribute to autoimmunity, as seen in an autoimmune animal model of systemic lupus erythematosus (Morris et al., 2019). B1 cells also produce granulocyte macrophage colony stimulating factor (GM-CSF), one of the growth factors that is increased in MDD (Maes et al., 2022) and which may further activate the production of IgM (Weber et al., 2014; Zasada et al., 2016). Finally, when triggered by bacterial antigens by ligation of their Toll-Like Receptors or the B cell receptor (BCR), B cells egress to the spleen and develop into plasma cells secreting cytokines and pathogen-specific IgM (Savage and Baumgarth, 2015; Rahman et al., 2016; Baumgarth et al., 2015). It should be added that in some conditions, CB2 ligation with agonists may enhance inflammatory responses (Karsak et al., 2007). Furthermore, 2-AG may stimulate the secretion of chemokines that enhance leukocyte adhesion and recruitment, and the production of TNF-α by endothelial cells (Atalay et al., 2019; Jehle et al., 2021; Gasperi et al., 2014). CB2 may act as a costimulator in mitogen-activated kinase (MAPK) and stress-activated kinase (SAPK) pathways, which promote mitosis, cell survival, and differentiation (Carayon et al., 1998).

Overall, the T cell activation in MDD may have stimulated B cells to acquire more CB2, which, in conjunction with the increased bacterial translocation in MDD, may result in an overproduction of IgM antibodies, which may have a variety of deleterious effects that exacerbate the primary T cell activation and thus the severity of the illness’s phenome. Moreover, increased CB2 may contribute to increased bacterial load in the systemic circulation due to increased production of IL-10 (Csoka et al., 2009).

### Effects of CBD

The fourth major finding of this study is that CBD concentrations between 0.1 and 1 µg/dL (therapeutic levels) had no discernible effect on any of the stimulated lymphocyte populations evaluated in this study. Previously, we did not observe any effect of CBD on the elevated levels of pro-inflammatory markers (M1, Th-1, Th-17, IRS, and T cell proliferation) and cytokines in MDD patients or healthy controls (Rachayon et al., 2022). These results demonstrate that CBD does not normalize the abnormalities in the prevalence of CD20+CB2+ populations and does not ameliorate the deficiencies in CD3+/CD4+CB2+ populations that predict the phenome of depression. CBD has a lower affinity for CB1 and CB2 compared to other cannabinoids such as AEA, 2-AG, and THC, as described in the Introduction. CBD is a partial agonist at CB2, which may at least partially mediate CBD’s anti-inflammatory effects (An et al., 2020). In addition, CBD desensitizes TRPV1 receptors, which have inflammatory activities, has agonist activities at TRPA1 receptors, which promote inflammation, depression, and anxiety, and acts as an antagonist at TRPM8 receptors, which reduce inflammation (Muller et al., 2019; An et al., 2020).

In addition, the higher CBD levels of 10 µg/dL substantially increase the stimulated levels of CD20+/CD3+/CD+4/CD8+ CB2+ cells following anti-CD3/CD28 treatment. Previously, we reported that 10 µg/dL CBD had significant effects on the immune system, inhibiting CIRS functions such as the stimulated production of IL-10, IL-13, and sIL-1RA, and IRS profiles, such as Th-1 and Th-17 (Rachayon et al., 2022). Consequently, higher CBD doses may have detrimental effects.

## Limitations

This study would have been more interesting if we had measured CB2 on other immune populations, including macrophages, dendritic cells, and natural killer cells. Future research should investigate the effects of CBD and other cannabinoids on the stimulated number and expression of CB2+ immunocytes. It could be argued that the sample size is rather small, although the minimal sample size was determined using power analysis. Furthermore, post-hoc power analysis performed on the primary analysis (regression of phenome on immune cell populations) shows that the actual obtained power is 0.98.

## Conclusions

There was an inverse relationship between the number of reduced CD4+CB2+ cells and M1, Th-1 and Th-17 phenotypes. MDD is characterized by a reduced FoxP3+CB1+ percentage and a higher CD20+CB2+ percentage. CD20+CB2+% (positively) and CD3+CB2+% (negatively) explained 33.2% of the variance in the depression phenome which comprises severity of depression, anxiety, and current suicidal behaviors. Decreases in FoxP3+CB1+% and CD3+/CD4+CB2+% contribute to immune homeostasis deficits in MDD, whereas an increase in CD20+CB2+% may contribute to the pathogenesis of MDD by activating T-independent humoral immunity. CB1 on Treg cells and CB2 on CD3/CD4+ and CD20+ cells are new drug targets to treat MDD.

## Author’s contributions

Design of the study: MM and MR. Recruitment of the participants: MR and KJ. Assays: PS. Statistical analyses: MM. Visualization: MM and PS. First draft: MM. Editing: MR, KJ, AS, AFA, and PS. All authors agreed to publish the final version of the manuscript.

## Ethical statement

All subjects gave their informed consent for inclusion before they participated in the study. The study was conducted in accordance with the Declaration of Helsinki of 1975, revised in 2013, and the protocol was approved by the Ethics Committee of the Institutional Review Board of the Faculty of Medicine, Chulalongkorn University, Bangkok, Thailand (#528/63)

## Data Availability Statement

The dataset generated during and/or analyzed during the current study will be available from the corresponding author (M.M.) upon reasonable request and once the dataset has been fully exploited by the authors

## ELECTRONIC SUPPLEMENTARY FILE (ESF)

**ESF, Figure 1.**
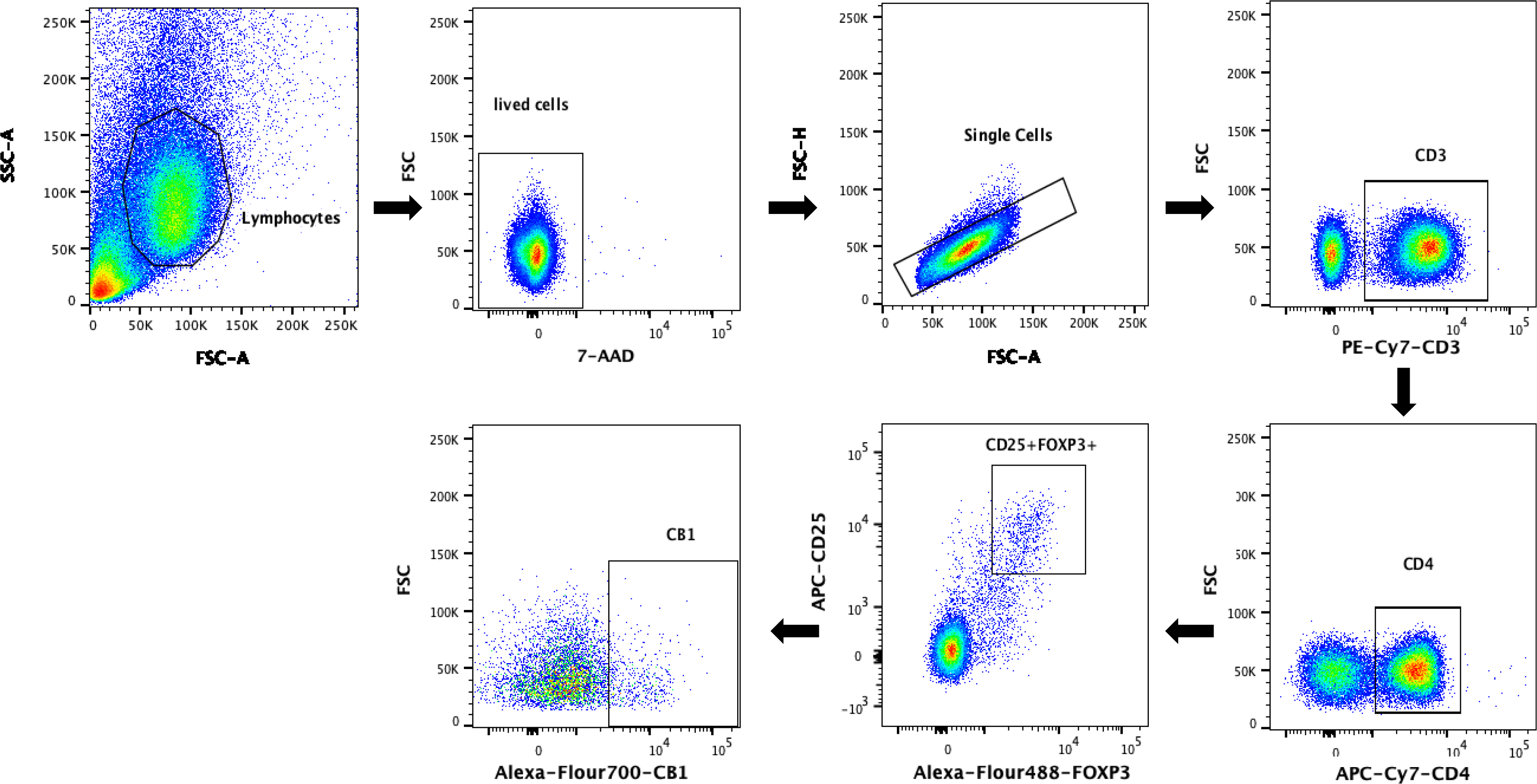
Gating strategy to assess FoxP3+CB1+ cells

**ESF, Figure 2.**
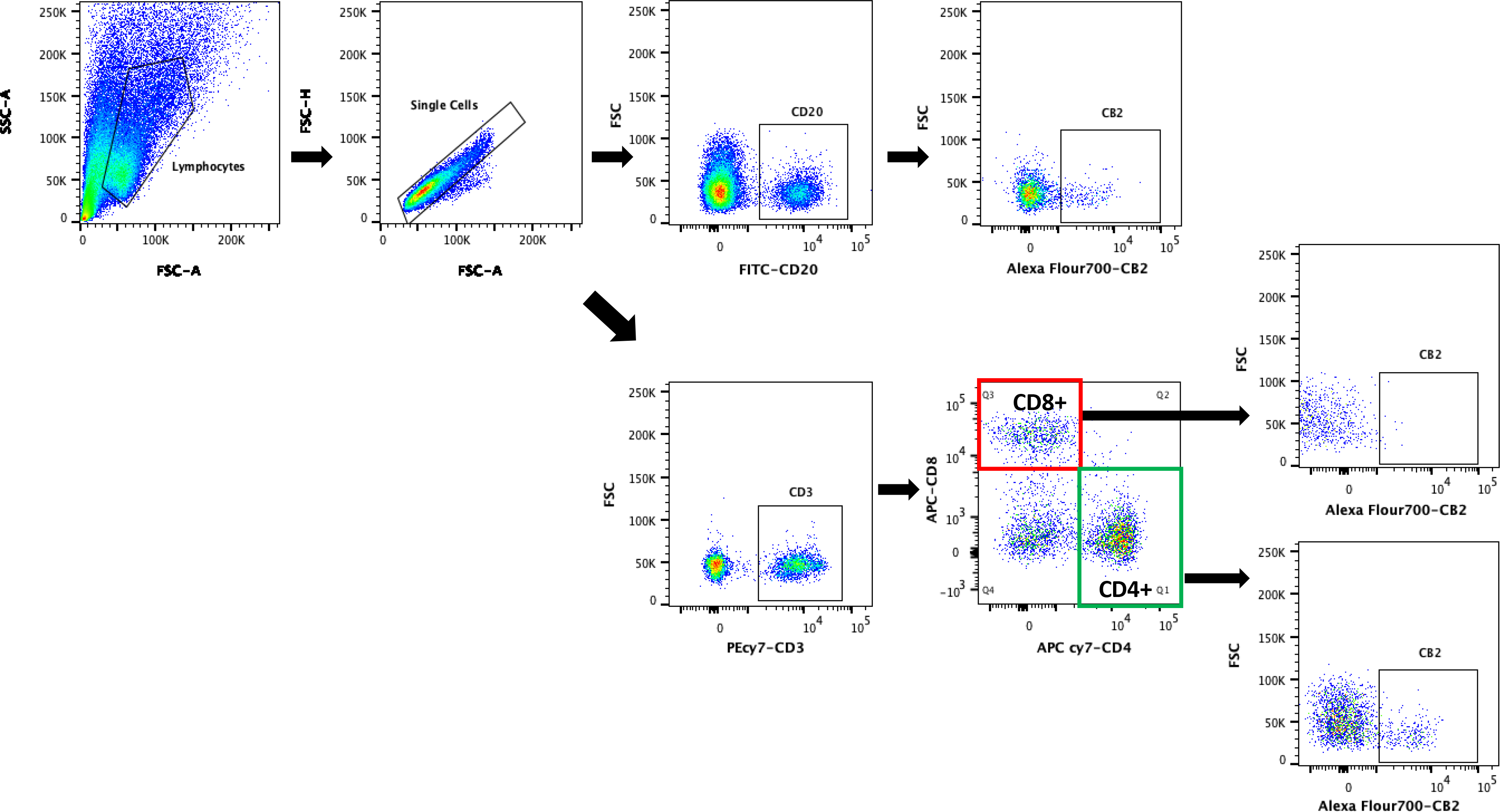
Gating strategy to assess CD4+/CD8+/CD20+ CB2+ cells

**ESF Table 1.** Description of the immune profiles used in this study

| Immune Profile | Members |
| --- | --- |
| M1 macrophage | IL-1β, sIL-1RA, IL-6, TNF-α, CXCL8, CCL3 |
| T helper-1 | IL-2, IFN-γ, IL-12 |
| T helper-17 | IL-6, IL-17 |
| IRS | IL-1β, IL-6, TNF-α, CXCL8, CCL3, IL-2, IFN-γ, IL-12, IL-17, IL-15, G-CSF, GM-CSF, CXCL10, CCL5, CCL2 |
| T cell growth | IL-4, IL-9, IL-12, GM-CSF, IL-15 |

IRS: immune-inflammatory response system; CIRS: compensatory immunoregulatory system
**ESF, Table 2** Effects of three concentrations of CBD on the anti-CD3/CD28 stimulated FoxP3+CB1+ cells.

**ESF, Table 2.** Effects of three concentrations of CBD on the anti-CD3/CD28 stimulated FoxP3+CB1+ cells.

| Estimates |  |  |  |  |
| --- | --- | --- | --- | --- |
| Treatment | Mean | Std. Error | 95% Wald Confidence Interval |  |
|  |  |  | Lower | Upper |
| STIM | 3.5060 | .26100 | 2.9944 | 4.0176 |
| CBDl | 3.5761 | .24482 | 3.0963 | 4.0560 |
| CBDm | 3.6179 | .25539 | 3.1174 | 4.1185 |
| CBDh | 6.2331 | .59754 | 5.0619 | 7.4042 |

Stim: stimulated with anti-CD3/CD28 beads, CBD (cannabidiol): all stimulated with anti-CD3/CD28 and 0.1 µg/dL CBD (CBDl), 1.0 µg/dL (CBDm) or 10.0 µg/dL (CBDh).

